# Clustering Countries on Development Indicators Reveals Structure Relevant for H5N1 Mortality Analysis

**DOI:** 10.1101/2025.11.08.25339808

**Authors:** Mwandida Kamba Afuleni, Leonardo Gada, Thomas House, Thomas Finnie

**Author notes:** Contribution considered equal (Mwandida Kamba Afuleni, Leonardo Gada).

## Abstract

Infectious diseases are often observed to have different epidemiology in different countries, which arises due to various factors including those that are ecological, socioeconomic, and healthcare-related. Such variability can sometimes be best captured through looking at groups of countries that are similar within-group but variable between-group. In this study we use statistical learning methods to generate data-driven disease-centric groupings of countries rather than those developed for administrative or political reasons by e.g. the WHO, World Bank, and the United Nations. In particular, we apply hierarchical clustering to group countries based on shared disease-relevant characteristics for zoonotic H5N1 influenza. Using statistical methods such as classification and regression trees (CART)-based imputation and dynamic tree cutting, the analysis accounts for missing data and identifies epidemiologically (rather than politically or economically) meaningful clusters. Applying health metric relevant indicators, we cluster the countries of the world and using a Bayesian approach compute CFRs of zoonotic H5N1 influenza before comparing across clusters. We find that countries with stronger healthcare systems and lower poverty rates tend to have lower and more stable CFRs, whereas resource-limited settings face higher fatality risks.

## 1 Introduction

Epidemiological analyses often rely on political or administrative boundaries and groupings to categorise countries when estimating disease metrics such as the case fatality ratio (CFR). However, infectious diseases are not believed to have their epidemiological characteristics like rate of spread and virulance determined by political and administrative factors; instead, we expect a much larger role for ecological, socioeconomic, and healthcare-related factors. Traditional groupings such as those defined by the World Health Organization (WHO), the World Bank, and the United Nations, may not always capture the most epidemiologically relevant distinctions for understanding disease burden and transmission risks [1, 2]. An alternative approach is to cluster countries based on shared disease-relevant characteristics, which may offer more meaningful insights for estimating key epidemiological quantities, particularly for emerging infectious diseases like zoonotic H5N1 influenza.

In this study, we apply hierarchical clustering, a well-established class of statistical methods widely used in various fields, including epidemiology, to identify natural country groupings relevant to health metrics within complex datasets [3]. This method is readily available in statistical software such as R [4] and enables us to group countries with comparable disease risk profiles, improving CFR estimation and other epidemiological metrics. To enhance the robustness of our clustering approach, we integrate the classification and regression trees (CART)-based missing data imputation [5] and dynamic tree cutting [6]. Missing data, a common challenge in epidemiological datasets, is addressed using the multiple imputation by chained equations (MICE) package [5], ensuring reliable estimates despite incomplete records. Additionally, we employ the Dynamic Tree Cut algorithm [6] from the `dynamicTreeCut` R package, which allows for adaptive cluster detection based on data structure rather than static thresholds. This approach improves cluster identification in heterogeneous global datasets.

Using this clustering framework, we estimate group-level CFRs for H5N1 and compare the results with traditional geographic classifications, such as WHO regions. By leveraging a data-driven methodology, we demonstrate how clustering countries by disease-relevant characteristics provides insights into disease severity and risk assessment on a global scale.

## 2 Materials and Methods

### 2.1 Study area

This study focuses on countries that have reported occurrences of H5N1 Avian influenza and aims to classify these countries using hierarchical clustering. The countries are: Australia, Azerbaijan, Bangladesh, Cambodia, Canada, Chile, China, Djibouti, Ecuador, Egypt, India, Indonesia, Iraq, Lao PDR, Myanmar, Nepal, Nigeria, Pakistan, Spain, Thailand, Turkey, United Kingdom, United States, and Viet Nam. We will sometimes refer to these as the “countries of interest” for our study.

### 2.2 Data sources

#### 2.2.1 Confirmed cases and deaths

Reported cases and deaths of H5N1 beginning from 2003 to 2024 were sourced from a publicly available database, the World Health Organization (WHO) tracker [7].

#### 2.2.2 Countries and indicators relevant to the H5N1

Data on countries around the world and indicators associated with H5N1 were obtained from the World Bank website [8].

#### 2.2.3 Predefined clusters

The WHO classifies countries based on geographical regions and similarities of the health system. These regions include Western Pacific, South-East Asia, Africa, Eastern Mediterranean, European and Americas [9]. The World Bank classifies countries into several groupings, including clusters based on income levels, geographic regions, and lending eligibility. Among these, the most commonly used is the income-based classification, which groups countries into low-, lower-middle-, upper-middle-, and high-income economies according to gross national income per capita. This classification was used in our clustering analysis [10]. The United Nations, on the other hand, provides country groups for both statistical and political purposes. For our analysis, we adopted the statistical regional clusters as defined in the UN mannual number 49 (M49) standard, rather than the political clusters that are primarily used for voting and diplomatic representation. The M49 statistical clusters include regions such as Africa, Asia, Europe, Americas, and Oceania [11].

### 2.3 Data analysis

#### 2.3.1 Country clustering

World Development Indicators (WDI) relevant to H5N1 used for clustering are: Population Total, Current Health Expenditure (% GDP), GDP Per Capita, Multidimensional Poverty Head-count Ratio (UNDP) (% population), Poverty Headcount ratio at Societal Poverty Line (% population), Rural Population, Prevalence of HIV, total (% population ages 15-49), Incidence of tuberculosis (per 100,000 people), Diabetes Prevalence (% population ages 20-79), Hospital beds (per 1,000 people), Physicians (per 1,000 people), Death rate, Crude (per 1,000 people), Rural land area (sq. Km) and Surface area (sq. Km) [8].

The data were cleaned by replacing e.g. missing or non-numerical values with `NA`. We ensured the numerical consistency of variables by converting relevant columns from character to numeric types. To reduce multicollinearity, we analysed pairwise correlations using Pearson’s correlation coefficient. Variables exhibiting high correlations (|*r*| > 0.80) were selectively removed while retaining the most informative variable. Specifically: Population Rural was retained while Population Total was removed. UNDP Multidimensional Poverty Headcount Ratio was retained while Poverty Ratio at Societal Poverty Line was removed. Rural Land Area was retained while Surface Area was removed.

We used the Multivariate Imputation by Chained Equations (MICE) package [5] in `R` to address missing data. The CART method was applied for data imputation [12, 13, 5] because, among the many available methods, it performs well with high-dimensional data, efficiently identifies important predictors, and naturally captures complex, non-linear relationships and interactions without requiring explicit specification. Imputation diagnostics, including strip plots and iteration trend plots, were evaluated to ensure the reliability of the imputed values. Prior to clustering, we standardised the variables using Z-score normalisation, where for a series of variables (*x*_*i*_) with mean *µ* and standard deviation *σ* we define a new series (*z*_*i*_) with

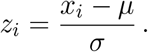

This step ensures equal weighting of variables during distance calculations.

We performed hierarchical clustering [3] to group countries into clusters based on their similarity. Clustering was conducted using the Manhattan distance metric [14, 15] and the complete linkage method [16, 3]. The Manhattan distance, defined as the sum of absolute differences between coordinates, was chosen for its robustness to outliers and its intuitive interpretability. This metric is particularly suitable for high-dimensional data. The number of clusters was determined using dynamic tree cutting with the `dynamicTreeCut R` package [6]. This method better accounts for the actual structure of the dendrogram, providing a more adaptive and data-driven identification of clusters. It enables the detection of subclusters of varying sizes and shapes, which was particularly important for our analysis. We chose dynamic pruning to explore the finer structure of the dendrogram and avoid an overly simplistic interpretation of the clustering that might result from applying a fixed, static cut.

#### 2.3.2 Case fatality rate (CFR)

Following clustering, we identified the clusters to which the countries of interest belonged and then computed the case fatality ratio (CFR) for H5N1 within each cluster using the methods described by Dunbar and Finnie (2021) [17, 18]. These methods follow a Bayesian approach. Total cases and deaths for each cluster were calculated, and the CFR was estimated using a Beta distribution. Uncertainty was quantified by calculating credible intervals (CrI). The Bayesian framework leverages conjugacy of the binomial likelihood with a beta prior on CFR – with the uninformative uniform prior we use a special case of the beta – to ensure stable estimates, particularly for smaller sample sizes. The output presents the CFR as a percentage with a 95% CrI, providing a more informative measure of disease severity.

## 3 Results

### 3.1 Clusters generated

Four clusters resulted from the analysis. The results of the whole world clustering including these 24 countries are presented in Table 1.

**Table 1.**
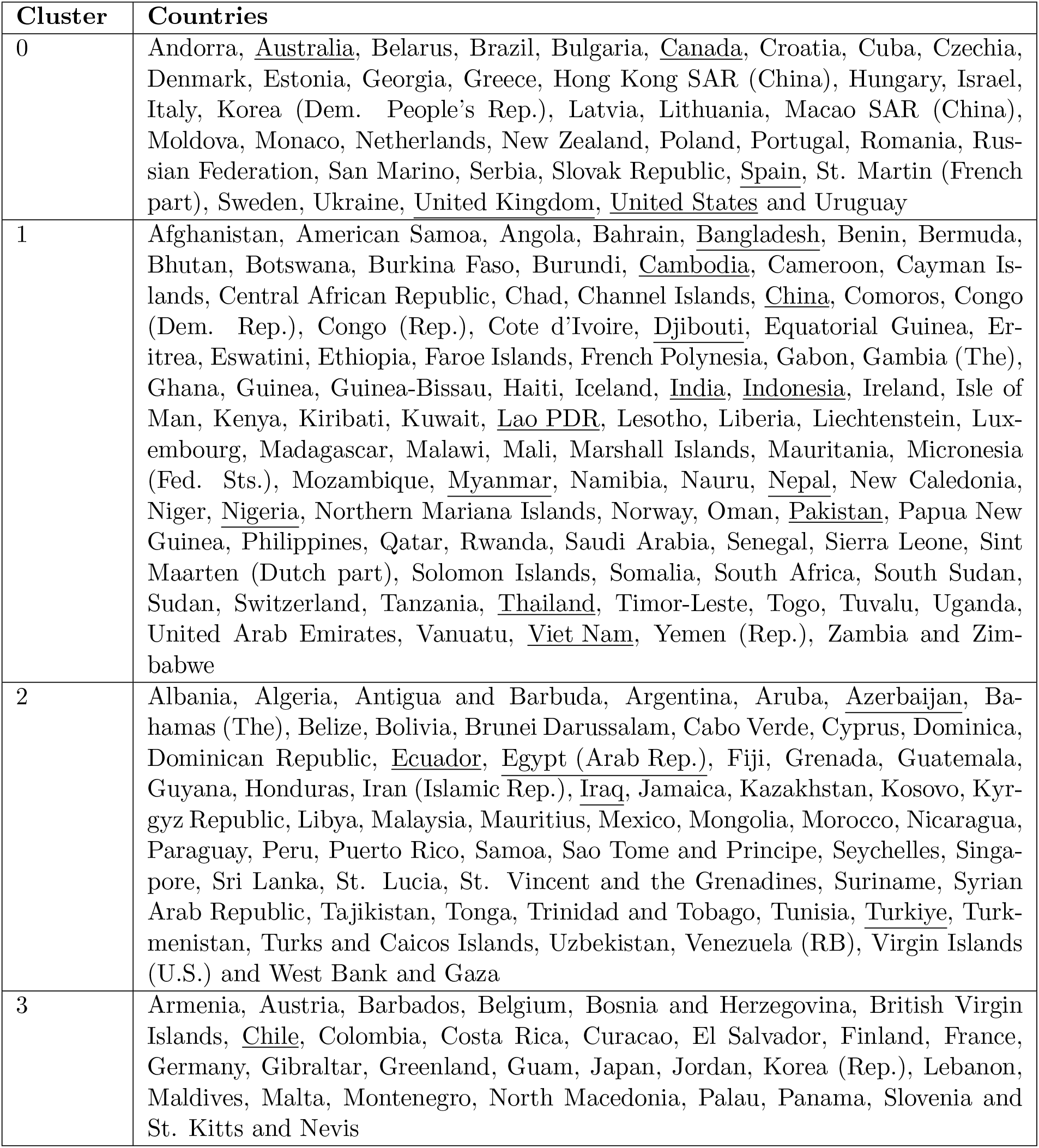
Results of data-driven country clustering. Countries of interest due to having experienced zoonotic H5N1 influenza cases in humans are underlined.

The four clusters were visualised on a world map, as shown in Fig. 1. The maps were generated using polygon data from the World Bank, openly available at [8]. Regions labeled as “NA” in the legend represent disputed territories or areas with missing polygon data (e.g. Western Sahara territories). Countries with similar geographic and epidemiological characteristics related to H5N1 have been grouped together.

**Figure 1.**
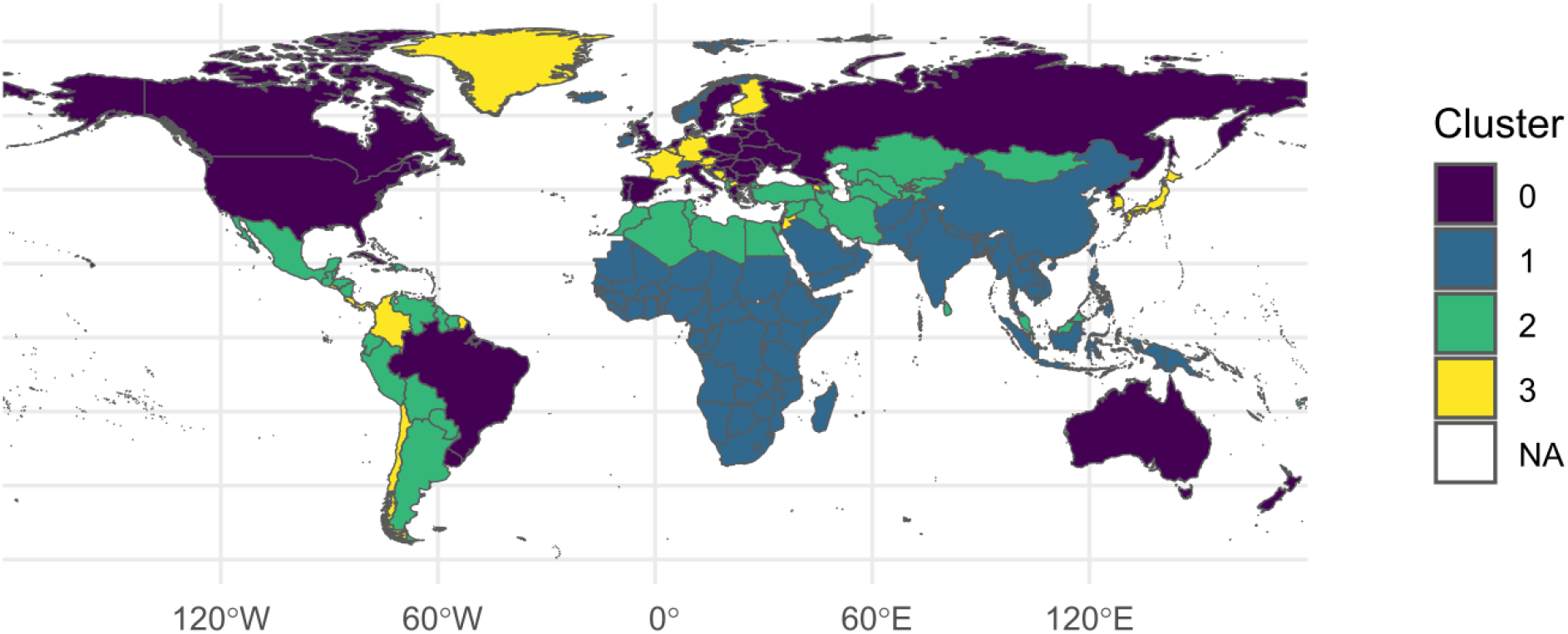
World map showing the clusters. The map was generated using the World Bank’s Official Boundaries dataset, available at: https://datacatalog.worldbank.org/search/dataset/0038272/World-Bank-Official-Boundaries. Disclaimer: The boundaries, colours, denominations, and any other information shown on this map do not imply, on the part of the authors, the journal, or the World Bank Group, any judgment on the legal status of any territory, or any endorsement or acceptance of such boundaries.

### 3.2 Existing clusters

Table 2 shows the grouping of the 24 countries by the World Health Organisation, the World Bank, and the United Nations. The United Nations categorises Turkey under Asia as well as Europe however, in this study, we have included it in Asia region, as the greater part of its landmass is geographically located in Asia rather than Europe.

**Table 2.**
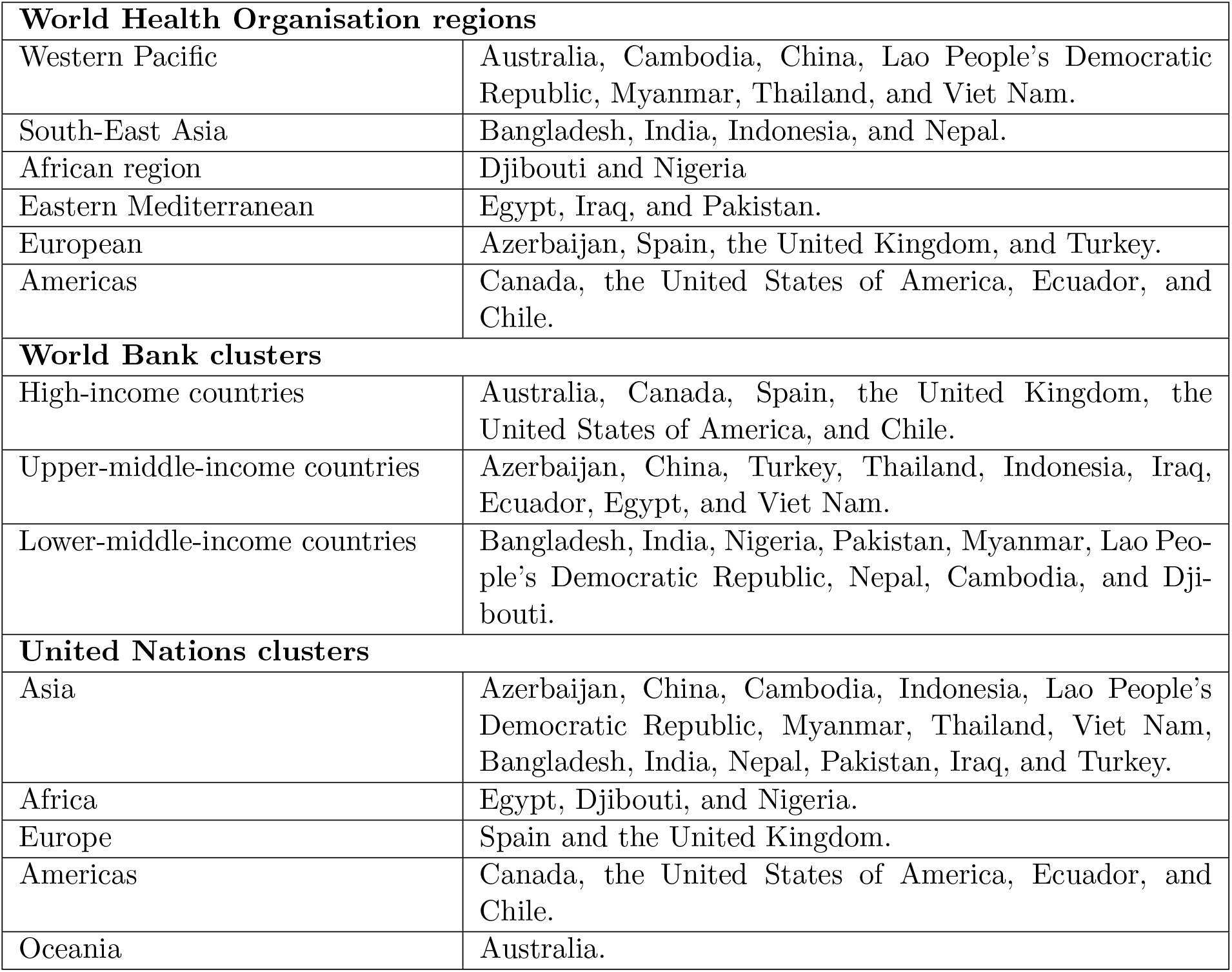
Groupings created for political and administrative reasons. The countries of interest are shown for the World Health Organisation, World Bank and United Nations clusterings

### 3.3 CFR for clusters

Fig. 2 illustrates CFR results for the clusters formed in this analysis as well as the CFR for the clusters formed by the WHO, World Bank and the UN. Cluster 0 (8.25%, CI: 1.23–24.87) had the lowest CFR however, its wide confidence interval suggests variability. Cluster 1 (66.22%, CI: 62.01–70.27) exhibited the highest and most stable CFR. Cluster 2 (34.26%, CI: 29.63–39.10) showed moderate and consistent CFRs, while Cluster 3 (29.29%, CI: 1.26–84.19) displayed extreme variability.

**Figure 2.**
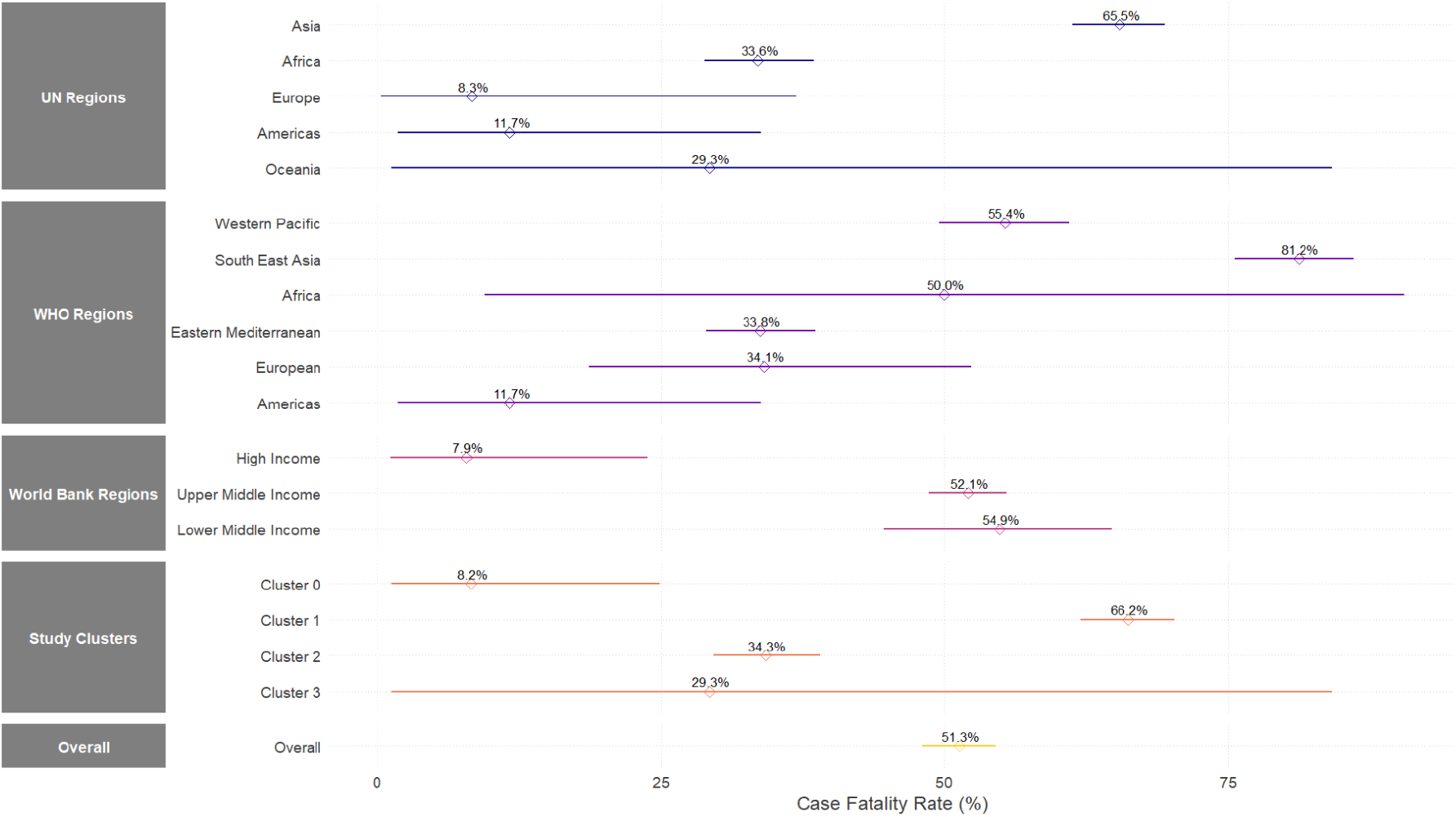
CFR of clusters and corresponding 95% credible interval (CrI)

For the WHO clusters, South-East Asia (81.23%, CI: 75.61–86.10) had the highest CFR, indicating severe outcomes and possible healthcare limitations. The Western Pacific (55.38%, CI: 49.61–61.05) and Africa (50.00%, CI: 9.43–90.57) also showed high CFRs, though Africa’s wide confidence interval suggests data variability. The Eastern Mediterranean (33.76%, CI: 29.04–38.70) and European (34.12%, CI: 18.64–52.35) regions had moderate CFRs, while the Americas (11.70%, CI: 1.78–33.87) reported the lowest.

According to World Bank clusters, High-income countries (7.86%, CI: 1.17–23.82) had the lowest CFR. Upper-middle-income countries (52.08%, CI: 48.60–55.54) and lower-middle-income countries (54.87%, CI: 44.70–64.78) presented significantly higher CFRs.

There are significant regional differences in H5N1 mortality for the United Nations clusters. Asia (65.48%, CI: 61.34–69.47) had the highest CFR, reflecting the heavy burden of H5N1 outbreaks in the region. Africa (33.58%, CI: 28.85–38.54) showed moderate CFRs, while Europe (8.30%, CI: 0.32–36.94) and the Americas (11.70%, CI: 1.78–33.87) had the lowest CFRs. Oceania (29.29%, CI: 1.26–84.19) displayed a wide confidence interval, suggesting uncertainty in CFR estimates.

### 3.4 Exploring Socioeconomic and Healthcare Disparities Across H5N1 Clusters

Selected key indicators of world development strongly linked to H5N1 were further analysed and compared across clusters formed in this study to provide deeper insights and explanations for the observed differences in CFR. These indicators include health expenditure, the number of physicians per 1,000 people, the number of hospital beds per 1,000 people, and the population proportion living in multidimensional poverty. Based on the box plot results found in the Appendices: Cluster 0 has the highest health expenditure, the most hospital beds and physicians per 1,000 people, and the lowest proportion of the population living in multidimensional poverty compared to the other clusters. Cluster 1 spends the least on health, has the lowest number of hospital beds and physicians per 1,000 people, and also experiences the highest level of multidimensional poverty. Clusters 2 and 3 fall in between, with moderate levels of these indicators.

Further analysis of the key world development indicators within the identified clusters revealed distinct patterns. Specifically, the cluster characterised by a high poverty ratio also showed significantly lower health expenditure, a smaller number of physicians per 1,000 people, and fewer hospital beds per 1,000 people, as illustrated in Fig. 3. Conversely, clusters with lower poverty ratios demonstrated better healthcare infrastructure and investment. Similarly, Fig. 4 highlights that the cluster with low GDP per capita was associated with higher death rates and a greater prevalence of both HIV and tuberculosis (TB). A cluster with higher GDP per capita, on the other hand, had lower disease prevalence however, the crude death rate was high.

**Figure 3.**
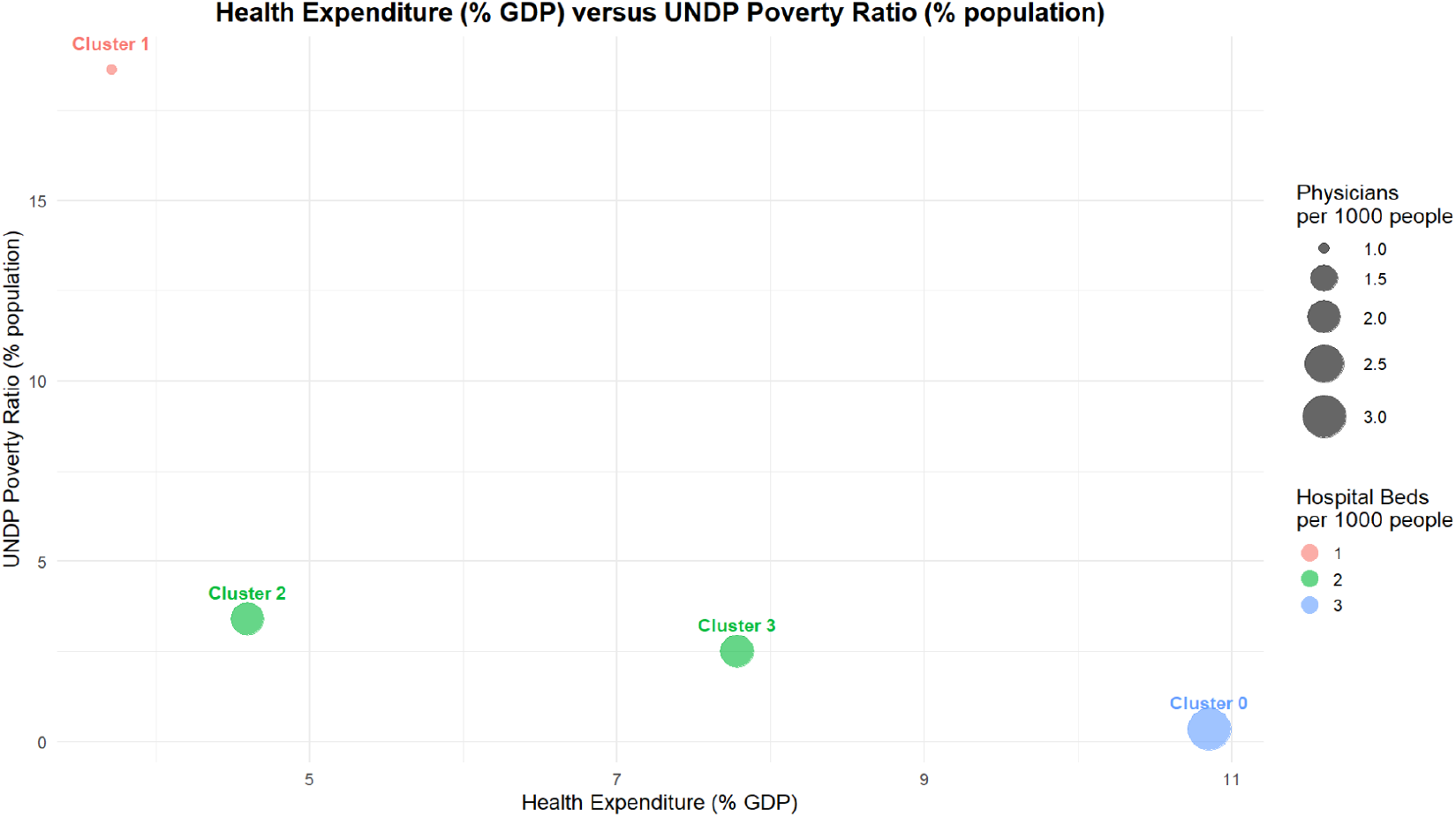
First group of selected health metrics for the four data-driven clusters.

**Figure 4.**
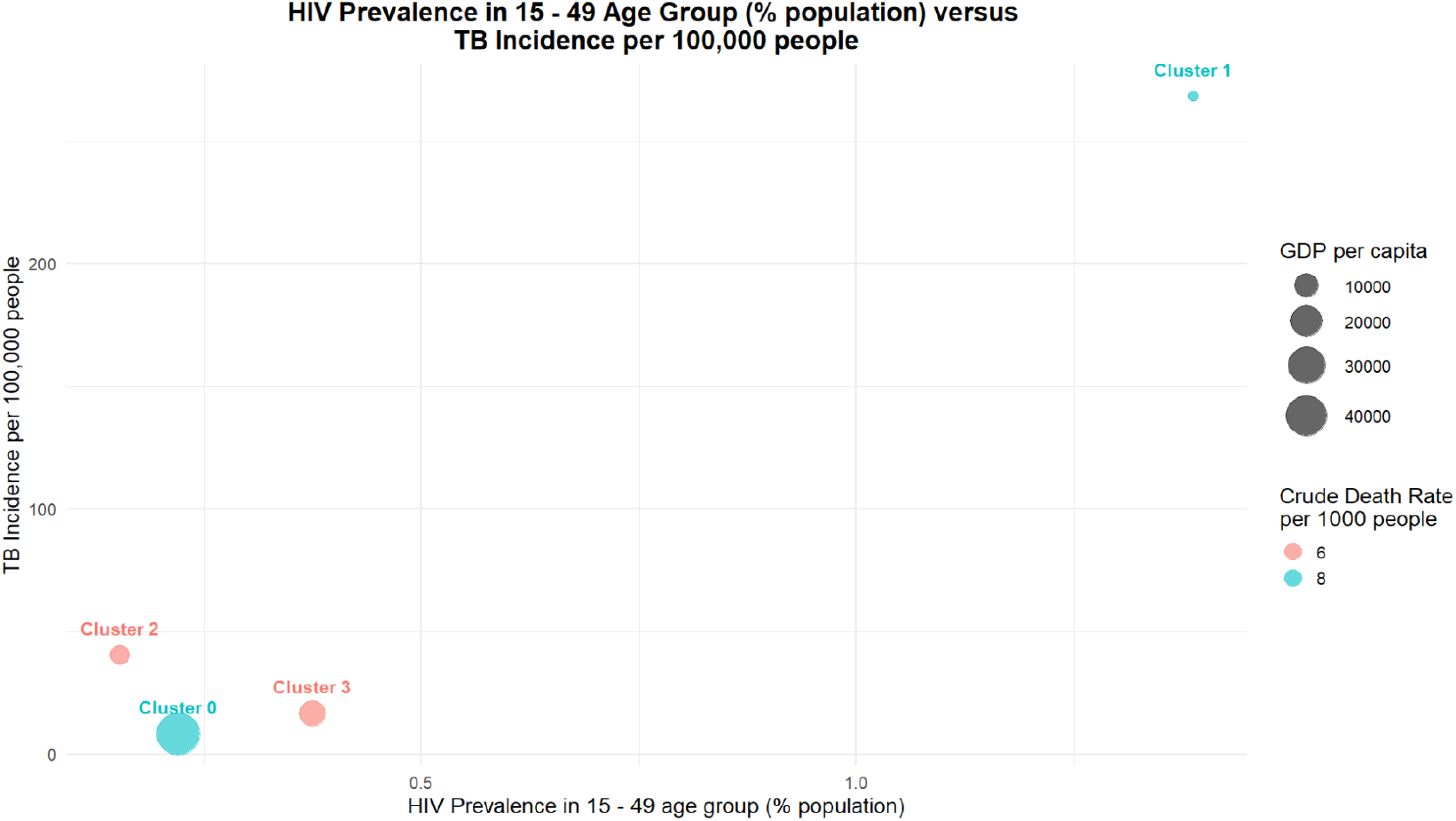
Second group of selected health metrics for the four data-driven clusters.

## 4 Discussion

In this study, we employed hierarchical clustering with dynamic tree cutting methods to group countries based on a set of disease-relevant characteristics. Our clustering approach incorporated key health and socioeconomic indicators. By utilising the CART data imputation method, we ensured that missing values did not bias the clustering process and that our analysis was based on a comprehensive dataset.

It is important to note that these clusters are not fixed or universally applicable. The classification presented here is derived from specific variables selected for this particular disease context. In this analysis, clustering was performed using indicators directly relevant to the metric of interest, which is the Case Fatality Ratio. Consequently, researchers focusing on other diseases or using different sets of indicators may generate entirely different cluster groupings. This reinforces the context-dependent nature of clustering results and highlights the necessity of tailoring clustering frameworks to the unique epidemiological, demographic, and health system attributes relevant to each research question.

Our results reveal that the clusters generated through this methodology differ significantly from pre-existing country classifications provided by organisations such as the World Health Organization (WHO), the World Bank, and the United Nations (UN). This divergence suggests that the selection of indicators plays a critical role in clustering countries when assessing disease burden and healthcare resource distribution. Traditional clustering frameworks may not adequately capture the multifaceted nature of disease prevalence and healthcare system capacities, highlighting the need for tailored clustering approaches that incorporate diverse and relevant indicators.

The observed differences in clustering patterns have important implications for global health policy and resource allocation. The reliance on conventional country groupings may overlook nuanced disparities in health-related factors that impact disease transmission, healthcare access, and overall health outcomes. Our approach demonstrates that hierarchical clustering with dynamic tree cutting provides an adaptable and data-driven means of categorising countries based on epidemiological and healthcare-related metrics. This method allows for the identification of more homogeneous groups, potentially improving the targeting of interventions and resource distribution strategies.

Cluster 0 had the lowest CFR (8.25%) but exhibited high variability, likely due to its strong healthcare infrastructure, including high health expenditure, more hospital beds, and a lower poverty rate. In contrast, Cluster 1 had the highest and most stable CFR (66.22%), aligning with its low healthcare investment, fewer medical resources, and high multidimensional poverty. Clusters 2 (34.26%) and 3 (29.29%) showed moderate CFRs, with Cluster 3 displaying extreme variability.

The CFR results from the clusters formed in our study show a broader range of values compared to the regional and income-based clusters from the WHO, World Bank, and UN. Cluster 1 in our study, with a CFR of 66.22% (62.01–70.27%), stands out with the highest mortality, which aligns more closely with the South-East Asia region (81.23%, CI: 75.61–86.10%) in the WHO clustering, suggesting that both groupings share characteristics of higher CFR most likely due to healthcare limitations or outbreak severity. On the other hand, Cluster 0 (8.25%, CI: 1.23–24.87%) from our study, with the lowest CFR, is more comparable to high-income countries in the World Bank clustering (7.86%, CI: 1.17–23.82%).The CFR results of Cluster 2 (34.26%, CI: 29.63–39.10%) and Cluster 3 (29.29%, CI: 1.26–84.19%) from our study are more in line with the African region (33.58%, CI: 28.85–38.54%) from the UN clustering, reflecting moderate outcomes but with notable variability, possibly due to differences in healthcare access and outbreak response. Overall, while there are similarities in CFR patterns between the clusters from our study and those formed by the WHO, World Bank, and UN, the detailed clusters in our study show more specific variations in CFRs, which may be influenced by regional, income, and epidemiological factors.

## 5 Conclusion

Our study demonstrates that hierarchical clustering with dynamic tree cutting provides a nuanced and data-driven approach to categorising countries based on disease-relevant characteristics. The divergence between our clustering results and traditional classifications by global organisations underscores the importance of selecting appropriate indicators when assessing disease burden and healthcare resource distribution. Our findings suggest that conventional frameworks may overlook critical epidemiological and healthcare disparities, whereas our method allows for more precise identification of homogeneous groups. The variations in CFR across our clusters suggest that better healthcare resources and lower poverty levels may contribute to lower and more stable CFRs, whereas resource-poor settings, combined with high disease prevalence, face significantly higher fatality risks. These insights highlight the importance of tailored clustering methodologies in informing disease-specific policies and intervention strategies, advancing beyond traditional geographic classifications to more precise, epidemiologically meaningful groupings.

## Code availability

Code associated with this analysis is publicly available at: https://github.com/thomasallanhouse/ClusteringCountries

## Data Availability Statement

All data used in this study are publicly available from reputable international databases. Reported H5N1 case and death data (2003–2024) were obtained from the World Health Organization (WHO) Global Influenza Tracker [7], available at WHO H5N1 Case Tracker (2003–2025). Country-level indicators were sourced from the World Bank Open Data repository [8], accessible at https://databank.worldbank.org/source/world-development-indicators.

Regional and income-based classifications were derived from the following publicly accessible sources: the WHO regional groupings [9] (https://www.who.int/about/who-we-are/regional-offices), the World Bank income classification system [10] (https://blogs.worldbank.org/en/opendata/world-bank-countryclassifications-by-income-level-for-2024-2025), and the United Nations M49 statistical regions [11] (https://unstats.un.org/unsd/methodology/m49/).

All data sources are cited in the manuscript and can be freely accessed for verification and further analysis.

## Supplementary figures

**Figure S1.**
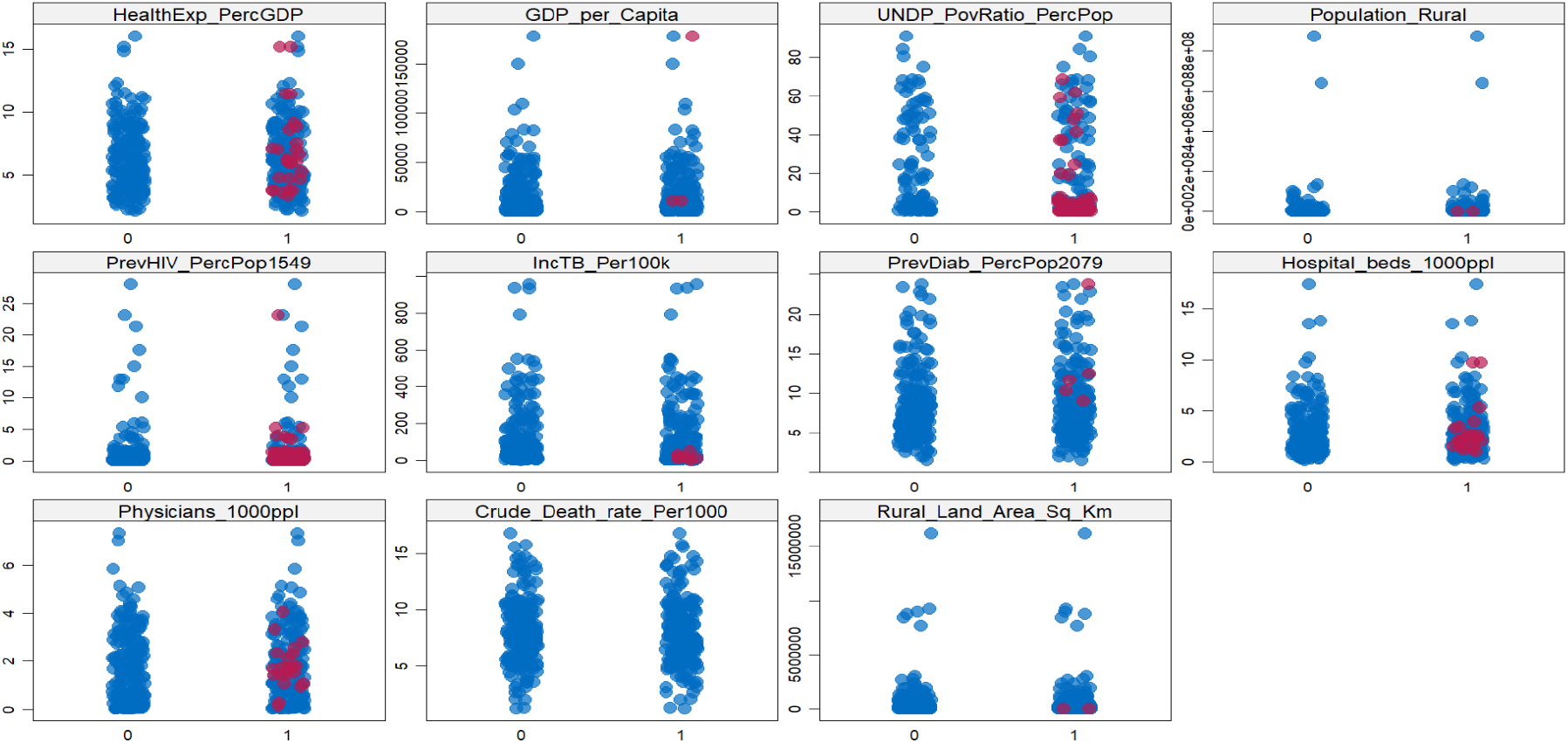
Strip plot diagnostics comparing imputed (red) and observed (blue) values for variables with missing data. The substantial overlap between red and blue points across all indicators indicates that the imputed values are consistent with the observed data, suggesting plausible imputations.

**Figure S2.**
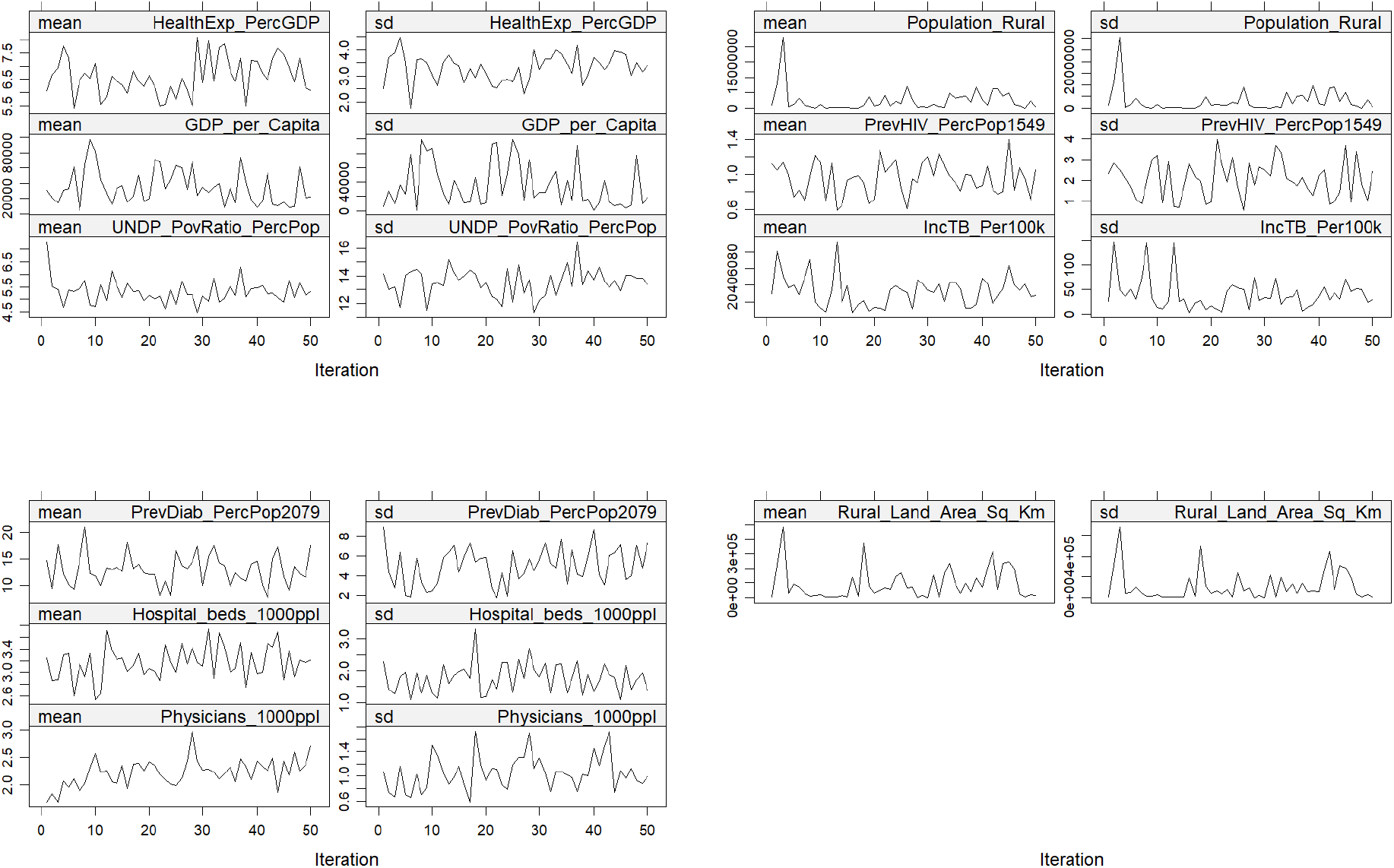
Iteration trace plots for convergence diagnostics, showing the mean and standard deviation of imputed variables across MICE iterations. The stabilisation of the lines indicates satisfactory convergence, suggesting that the imputed values are consistent across iterations for most indicators.

**Figure S3.**
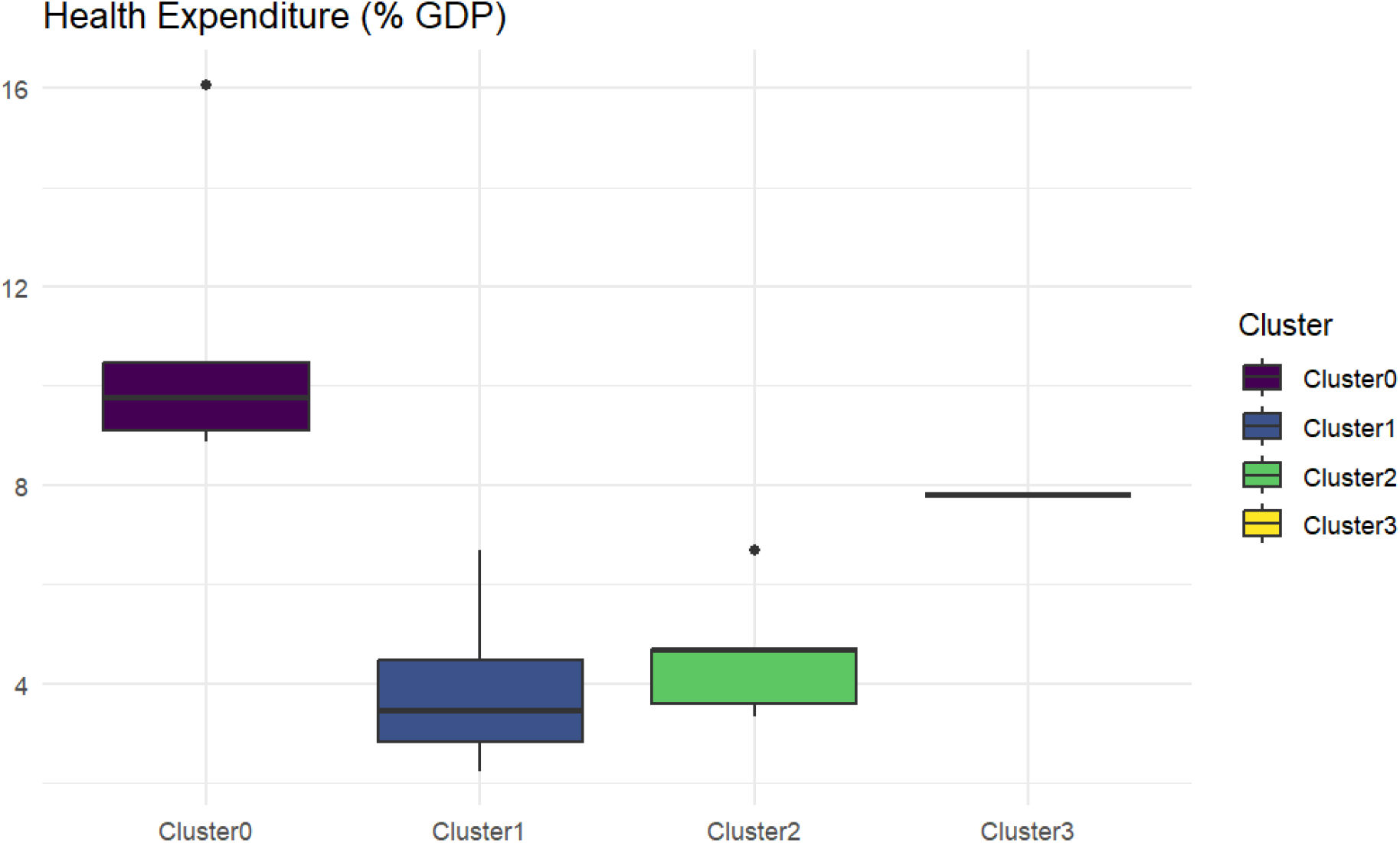
Boxplots comparing the distribution of health expenditure (% of GDP) among the four clusters.

**Figure S4.**
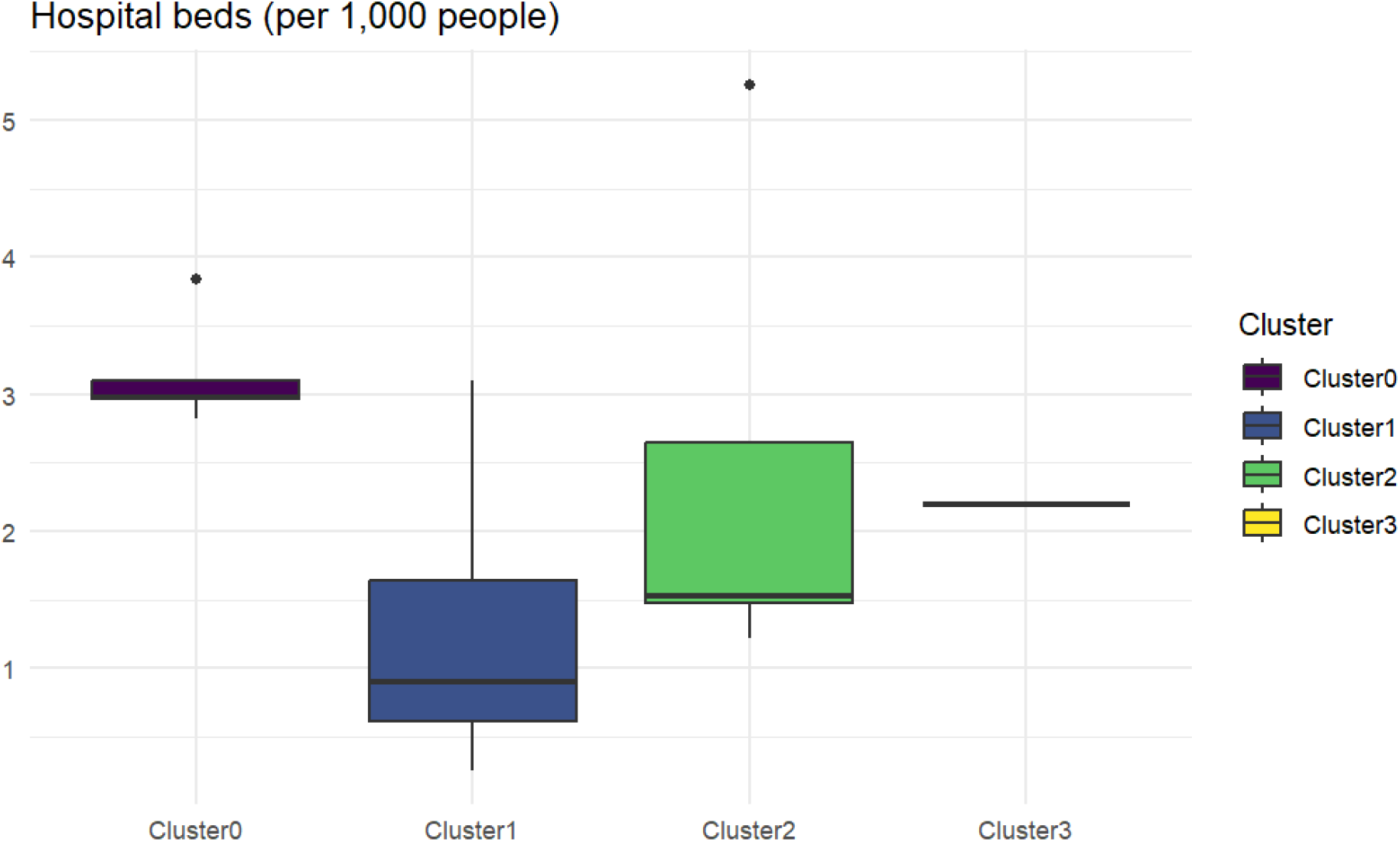
Boxplots comparing the distribution of the number of hospital beds (per 1,000 people) among the four clusters.

**Figure S5.**
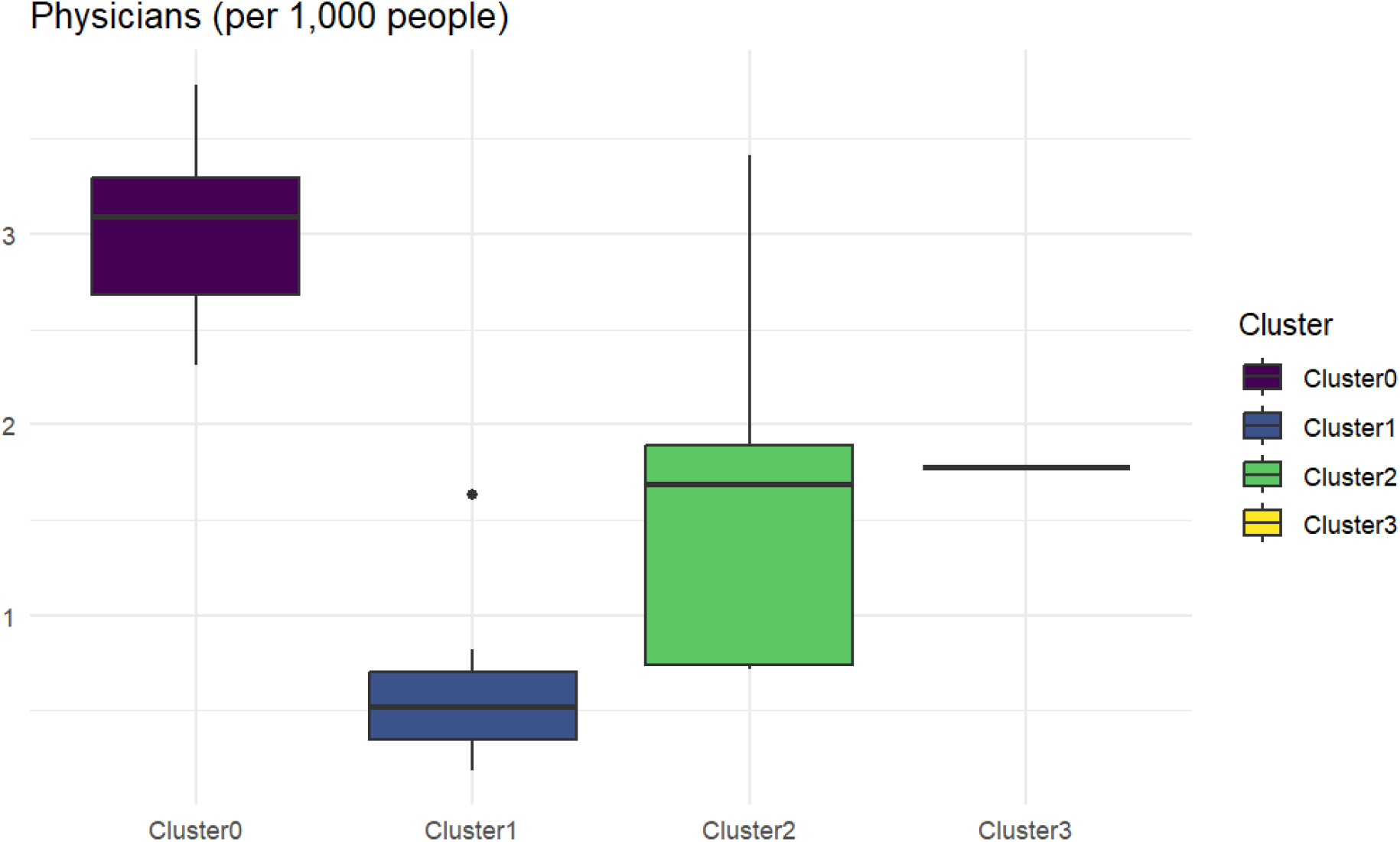
Boxplots comparing the distribution of the number of physicians (per 1,000 people) among the four clusters.

**Figure S6.**
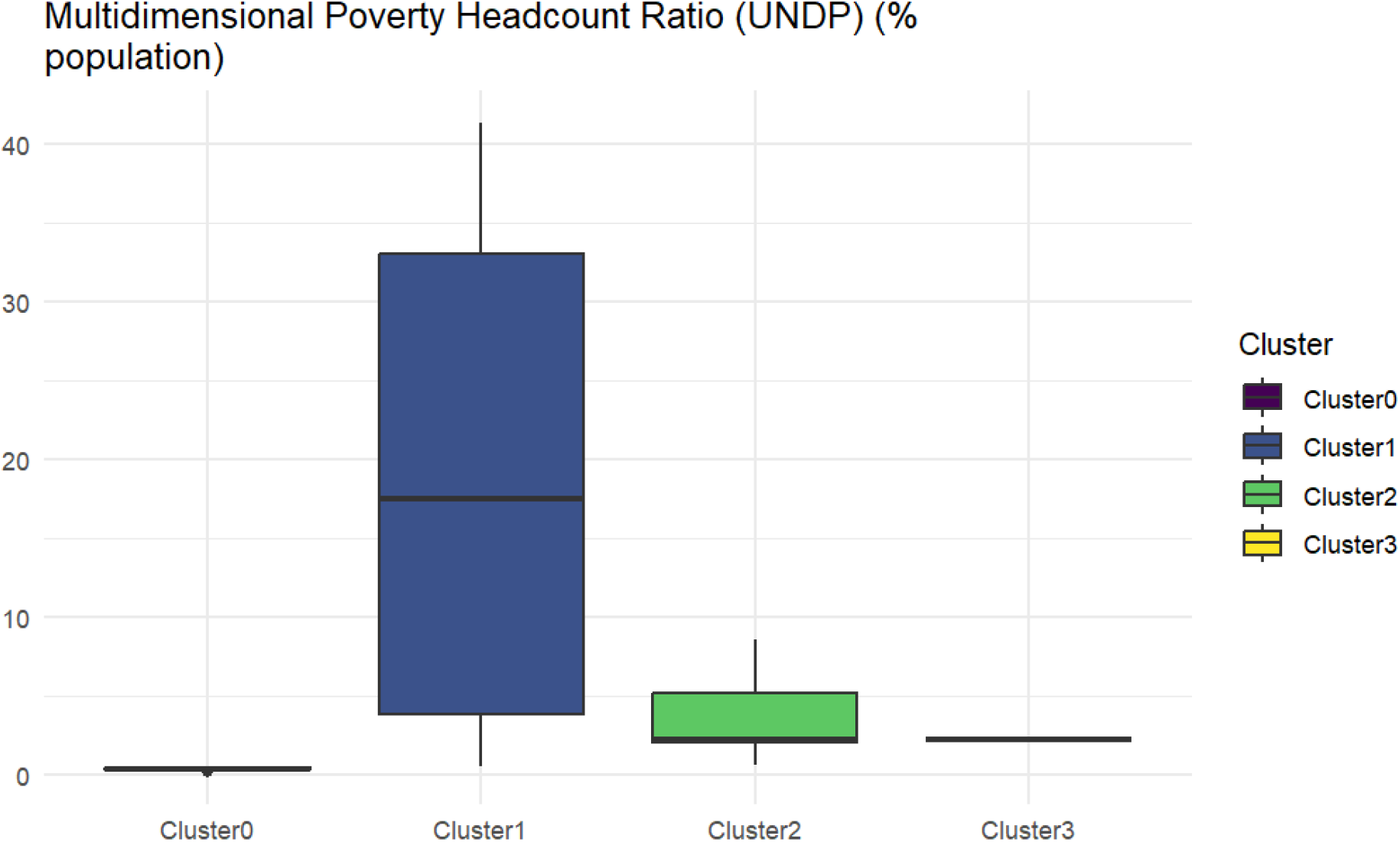
Boxplots comparing the distribution of UNDP poverty ratio (% of population) among the four clusters.

## References

1. Morens DM, Folkers GK, and Fauci AS. The challenge of emerging and re-emerging infectious diseases. Nature 2004; 430:242–9

2. Jones KE, Patel NG, Levy MA, Storeygard A, Balk D, Gittleman JL, and Daszak P. Global trends in emerging infectious diseases. Nature 2008; 451:990–3

3. Murtagh F. Ward’s Hierarchical Clustering Method: Clustering Criterion and Agglomerative Algorithm. ArXiv abs/1111.6285 2011

4. R Core Team. R: A Language and Environment for Statistical Computing. R Foundation for Statistical Computing. Vienna, Austria, 2021. Available from: https://www.R-project.org/

5. Van Buuren S and Groothuis-Oudshoorn K. mice: Multivariate imputation by chained equations in R. Journal of statistical software 2011; 45:1–67

6. Langfelder P, Zhang B, and Horvath S. Defining clusters from a hierarchical cluster tree: the Dynamic Tree Cut package for R. Bioinformatics 2008; 24:719–20

7. Organization WH. Global Health Observatory data repository. 2025. Available from: https://www.who.int/publications/m/item/cumulative-number-of-confirmed-human-cases-for-avian-influenza-a(h5n1)-reported-to-who--2003-2025--20-january-2025

8. Bank TW. World Development Indicators. 2025. Available from: https://databank.worldbank.org/source/world-development-indicators

9. World Health Organization. WHO regional offices. 2025. Available from: https://www.who.int/about/who-we-are/regional-offices

10. World Bank. World Bank country classifications by income level for 2024–2025. 2025. Available from: https://blogs.worldbank.org/en/opendata/world-bank-country-classifications-by-income-level-for-2024-2025

11. United Nations Statistics Division. Standard country or area codes for statistical use (M49). 2025. Available from: https://unstats.un.org/unsd/methodology/m49/

12. Breiman L, Friedman J, Olshen RA, and Stone CJ. Classification and regression trees. Routledge, 2017

13. Therneau T, Atkinson B, and Ripley B. rpart: Recursive Partitioning and Regression Trees. R package version 4.1-15. 2019

14. Krause E. Taxicab geometry: An adventure in non-Euclidean geometry, DoverPublications. com. 1987

15. Deza E, Deza MM, Deza MM, and Deza E. Encyclopedia of distances. Springer, 2009

16. RR S. A statiscal method for evaluating systematic relationships. Univ Kans sci bull 1958; 38:1409–38

17. Dunbar MBN and Finnie TJ. bayesint: a Python package for calculating Bayesian credible intervals of ratios of beta distributions. Journal of Open Research Software 2021; 9

18. Bekker-Nielsen Dunbar M, Finnie TJ, Sloane B, and Hall IM. Methods for calculating credible intervals for ratios of beta distributions with application to relative risks of death during the second plague pandemic. PLOS ONE 2019; 14:e0211633

